# Management capacity of healthcare facilities in Low- and Middle-Income Countries; A scoping review

**DOI:** 10.1101/2025.03.11.25323726

**Authors:** Harrison Ochieng, Anita Musiega, Benjamin Tsofa, Jacinta Nzinga, Edwine Barasa

## Abstract

How health facilities are managed determines their performance and health service delivery. Management capacity of health facilities comprises the competency of managers at the individual level and the management support and work environment in their institutions. Evidence shows this management capacity influences service delivery and performance of the facility. For LMICs, there are evidence gaps as existing evidence is scarce, varied in the assessment of management capacity of PHC facilities and report a measurement gap due to the scarcity of assessment tools contextualised to the LMIC PHC setting. Our review aims to address these gaps by mapping and summarising the existing literature on management capacity of PHC facilities in LMICs, its components and performance across these components, providing evidence on what needs to be improved for better service delivery. We used Arksey and O’Malley’s scoping review methodology. We searched PubMed, Scopus, Web of Science and Google Scholar and hand-checked reference lists. We synthesized findings using a thematic approach. We included 21 articles out of the 3867 articles gotten. Individual capacity consisted of managerial competencies grouped into seven groups: (1) communication and information management, (2) financial management and planning, (3) human resource, supportive and performance management, (4) community stakeholder and engagement, (5) target setting and problem solving, (6) leadership and (7) situational analysis. Institutional capacity included functional support systems grouped into; (1) availability of resources, (2) support to undertake duties and (3) clear roles and responsibilities. Gaps were prevalent across individual and institutional capacities. There were deficiencies in the managerial competencies of the managers and the functional support systems were not adequate. These negatively affected facility service delivery and performance. There is still a scarcity of studies hence more research is needed. Furthermore, interventions such as training and supportive supervision should be considered in improving the managerial competencies of managers.

## BACKGROUND

Primary Healthcare (PHC) facilities are important in the provision of facility-based PHC services and coordinating services at the community level [1]. These facilities are often the community’s initial point of contact with healthcare services, serving as the gateway to the entire healthcare system. From existing evidence, management capacity of these facilities influences the availability of medicine and equipment, efficiency in the use of resources, availability and quality of care among other service delivery goals [2–4]. These represent inputs, outputs and outcomes that are integral to PHC performance based on the World Health Organization (WHO) PHC measurement framework [5]. Management capacity is also important for overall health system leadership and governance which strongly influences the achievement of health system goals [6].

Management capacity in PHC comprises the competency of managers at the individual level and the management support and work environment in their institutions. The United Nations Development Program (UNDP) defined capacity as the ability of individuals, institutions and societies to solve problems, perform functions and sustainably achieve set goals [7]. Applying this definition to health systems, the individuals are the managers across the system while the institutional arrangements include the management structures and support systems [8] [8]. The WHO leadership and management framework highlights four key dimensions for good leadership and management capacity; appropriate competencies, adequate number of managers, functional support systems and enabling working environment [9,10]. Broadly categorized, managerial competencies fall at the individual manager’s level, while the other three dimensions operate at the institutional level, shaped by actors in higher authority within the system [11].

Good management capacity is integral for better quality of services, patient outcomes and facility performance. Studies conducted in high-income countries have shown an association between management capacity at the individual and institutional level, and the quality of care. In these studies, having a competent manager and a high performing management board was associated with better care processes, high quality of care and reduced risk of mortality for different cardiovascular and respiratory conditions [12,13]. Better care processes, quality of care and outcomes are contributors to good facility performance [5].

Emerging evidence from Low- and Middle-Income Countries (LMICs) show that good management capacity is associated with better availability of essential medicines and quality of services. From this evidence, facilities with good management capacity have better patient reported quality of care and trust [3,14]. Patients were also more satisfied and likely to return to the facility [3,14]. These facilities also have better availability of essential medicines and equipment [15,16]. This evidence shows the important role that management capacity plays in influencing the availability of essential resources, quality of services and patient outcomes, all of which contribute to facility performance. However, similar studies are still scarce with little to no evidence for a good number of LMIC PHC contexts. Summarising the existing evidence is important in providing information regarding the components and level of management capacity, how it influences service delivery across different contexts and cross-learning on what can be improved for better service delivery.

In these few studies conducted in LMICs, there is variation in the assessment of management capacity and a reported measurement gap. Drawn from the WHO leadership and management framework and global literature, management capacity comprises individual and institutional capacities [7–11]. Compared to high-income countries, few studies have been conducted in the LMIC setting to assess management capacity of PHC facilities. In these studies, there is heterogeneity in the assessment of management capacity. Some of these studies have assessed varying managerial competencies alone while others have also included institutional capacities in the assessment of management capacity [11,17]. However, the distinction between individual and institutional capacities emerges in the work of Heerdegen et al., 2020 and Adeniran et al., 2022 but not as clearly stated. The existing studies also report a measurement gap in LMICs due to limited tools contextualised to the PHC settings. For instance, the existing World Management Survey (WMS) is suited to high income PHC settings and is expensive and time consuming to implement in LMIC settings [18,19]. This measurement gap and lack of studies mean for some LMIC contexts the evidence is not available. However, summarising evidence from other LMICs which have similar PHC contexts can provide useful information and cross-learning on areas needing improvement for better service delivery and facility performance.

Considering the limited and varied nature of the evidence in LMICs and the relevance of management capacity on service delivery and performance, it is important to map and summarize the existing findings. Existing reviews summarizing evidence on management capacity of PHC facilities have predominantly been done in high-income countries [20,21]. To the best of our knowledge, no reviews currently summarize the number of studies assessing management capacity of PHC facilities in LMICs, the specific components of this management capacity and performance in each of these components. Our review aims to address this gap by mapping and summarising the existing literature on management capacity of PHC facilities in LMICs, its components and performance across these components. This will provide valuable insights into what constitutes management capacity and evidence-based gaps that policymakers and other actors can address for better service delivery. It will also lay the groundwork for further research on management capacity.

## METHODS

In this scoping review, we adopted Arksey and O’Malley’s framework in examining the components of management capacity of PHC facilities in LMICs [22]. The framework is one of earliest and most used methodological guidance for scoping reviews, allowing for a systematic process in the retrieval, synthesis and reporting of the evidence [23]. Additionally, the framework is also appropriate and accommodating of diverse collaborating teams [24]. We reported the results according to the PRISMA extension for scoping reviews (PRISMA-ScR). The protocol for this review was not registered prior.

### Eligibility criteria

We selected eligible studies using the Population, Concept, and Context (PCC) framework recommended for scoping reviews [25].

*Population*- We included studies focusing on the management in PHC facilities comprising the managers and management teams.

*Concept*- We included studies that defined and assessed management capacity as part of the study objectives. These studies assessed the individual competencies of managers, the institutional capacities in the form of functional support systems and enabling working environments, management processes and functions. They also detailed the evaluation tools, frameworks and models used.

*Context* - We included studies conducted in PHC facilities in LMICs.

*Types of sources of evidence* - We included peer-reviewed studies and studies published in English.

We excluded studies whose focus was not on PHC facility management, conducted in secondary care settings, from high-income countries and those not published in English.

### Information sources and search

Using a comprehensive search strategy, we searched for relevant literature in four databases, namely PubMed, Scopus, Web of Science and Google Scholar. We developed a search strategy based on the PCC framework [25]. We conducted an initial search in PubMed, added more search terms and Boolean operators based on our interaction with the search results and conducted a second search using the refined search strategy. We then modified the search terms based on the different databases, conducting the search in Scopus, Web of Science and Google Scholar. The most recent search was conducted on 21^st^ October 2024. The search strategy for PubMed is found in the supplemental file.

### Selection of sources of evidence

We uploaded all the references to Covidence software and removed duplicates [26]. We used Covidence to streamline the review process from screening, selection and charting. Title and abstract screening, full-text screening and selection were done by two independent reviewers with blinding based on the predefined eligibility criteria. Disagreements were resolved through discussion and a consensus was reached.

### Data charting and items

We developed a data charting and extraction form priori based on the PCC framework. The form (supplemental file S1) was piloted using four studies and revised accordingly by two independent reviewers. We extracted information on general study characteristics, components of management capacity, performance in the components and the influence on facility service delivery and performance. One of the reviewers extracted the data and another reviewer conducted quality checks of the extracted data. Where there were discrepancies, we discussed them with the other co-authors and a consensus was reached.

### Synthesis of Results

We summarised the characteristics of the studies which included the number, country and economic classification, year of publication and design of the studies. We then conducted an inductive thematic analysis of the findings based on the recommendations of Braun & Clarke, 2006, grouping similar components [27].

## RESULTS

### Selection of sources of evidence

Our search strategy yielded 3867 articles (**Fig 1**). In Google Scholar, we selected the first 200 results based on the recommendations of Haddaway et al., 2015 and Briscoe et al., 2023 on the lesser relevance of studies past the 200^th^ mark [28,29]. We got 12 more studies from hand-searching reference lists. We removed 582 duplicates using Covidence and 1 article manually. During title and abstract screening, we excluded 3159 articles as they did not meet the inclusion criteria (Fig 1). We further excluded 104 articles at full-text review, remaining with a final 21 articles which were included in the review.

**Fig 1:**
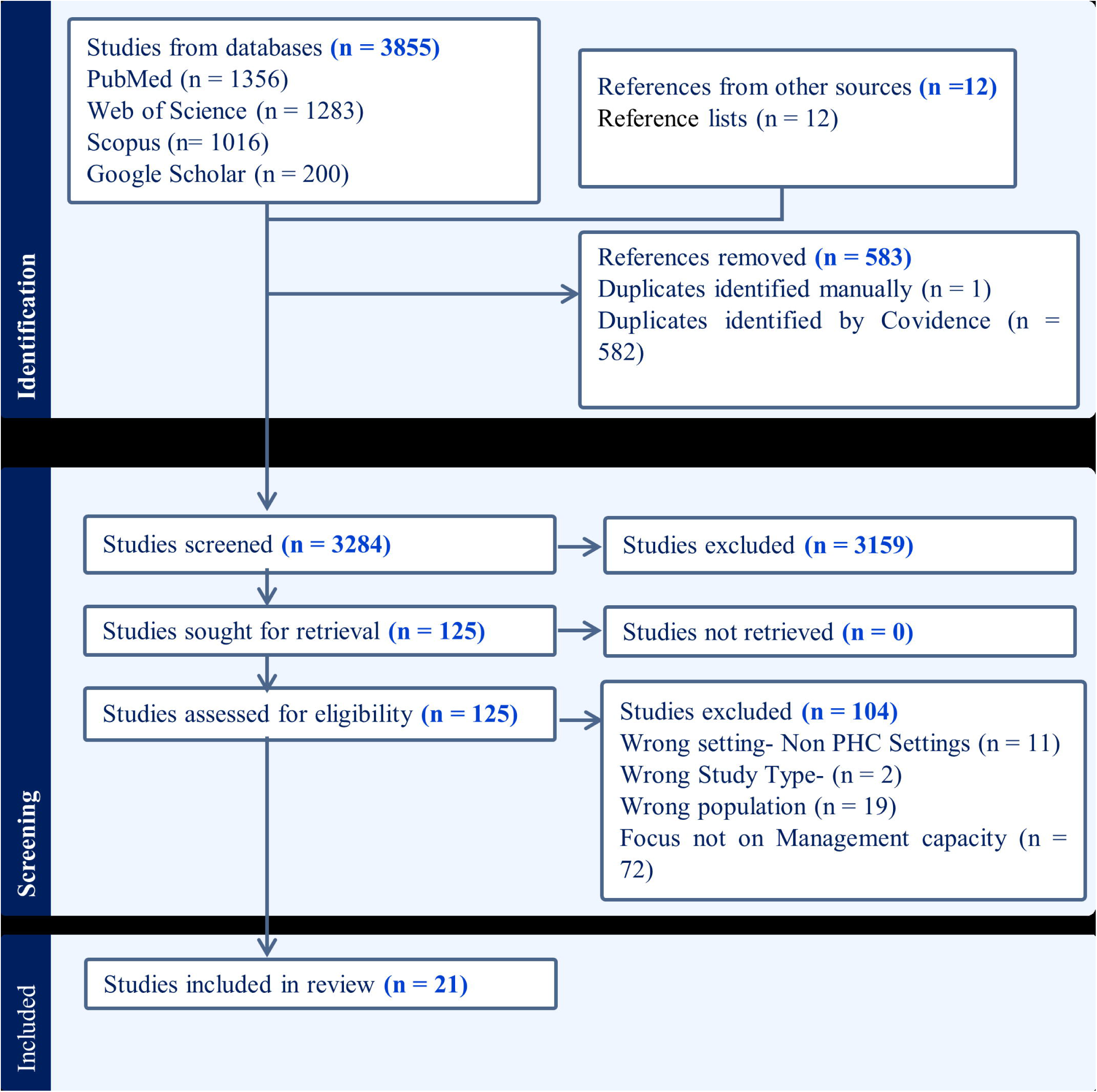
PRISMA Chart.

### Characteristics of sources of evidence

We synthesised 21 studies from 9 countries in Africa, 3 countries in Asia, 1 country in South America and 1 country in Europe (Table 1). The income levels varied from Low Income (LI), Lower Middle Income (LMI) to Upper Middle Income (UMI). The studies were published between 2012 and 2024. In terms of study designs, quantitative designs were the most common, used in 16 (76.2%) studies (Table 1). Other study included qualitative designs and mixed methods. A detailed list of the studies is available in the supplemental file.

**Table 1:**
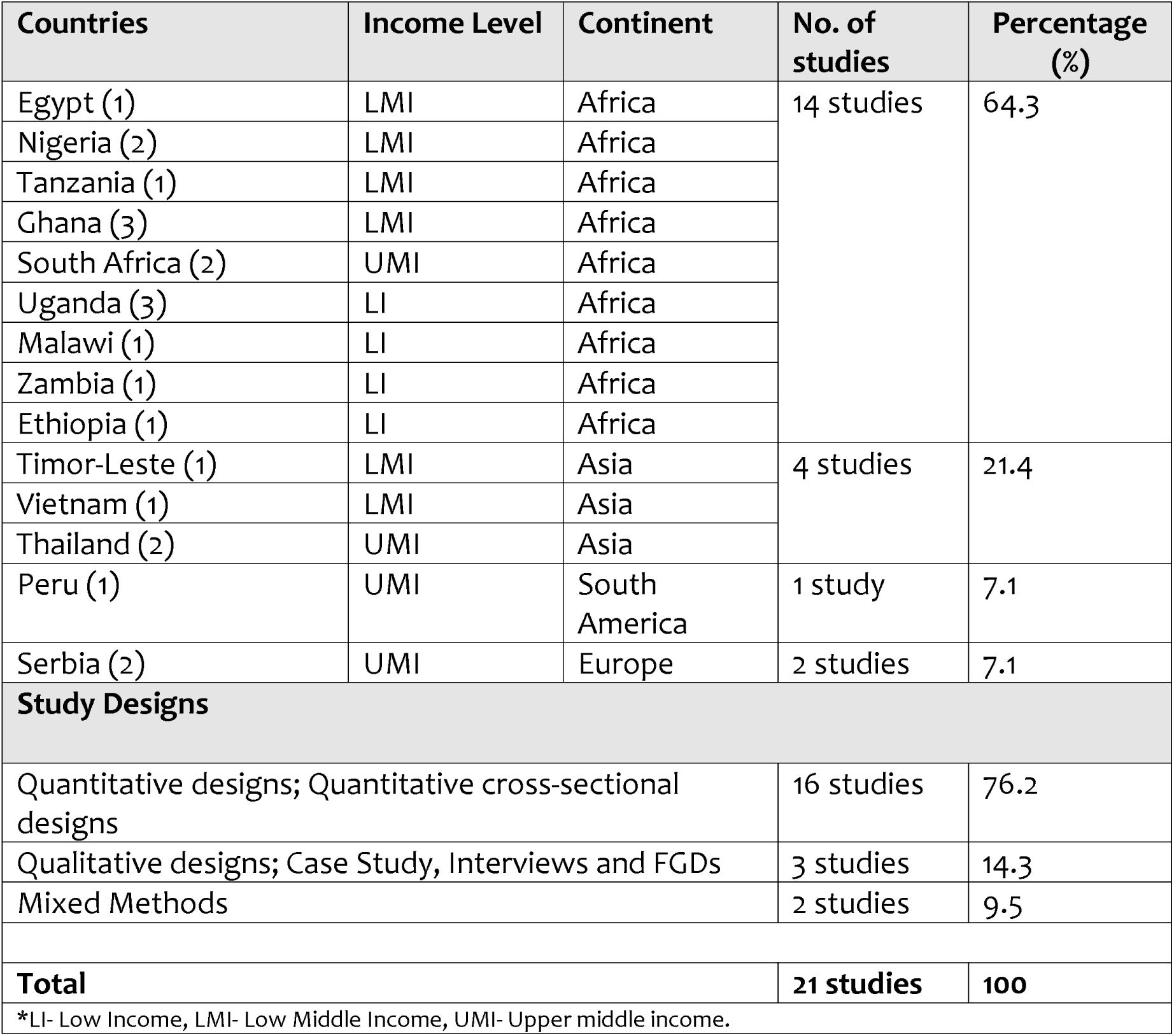
Characteristics of the studies.

| Countries | Income Level | Continent | No. of studies | Percentage (%) |
| --- | --- | --- | --- | --- |
| Egypt (1) | LMI | Africa | 14 studies | 64.3 |
| Nigeria (2) | LMI | Africa |  |  |
| Tanzania (1) | LMI | Africa |  |  |
| Ghana (3) | LMI | Africa |  |  |
| South Africa (2) | UMI | Africa |  |  |
| Uganda (3) | LI | Africa |  |  |
| Malawi (1) | LI | Africa |  |  |
| Zambia (1) | LI | Africa |  |  |
| Ethiopia (1) | LI | Africa |  |  |
| Timor-Leste (1) | LMI | Asia | 4 studies | 21.4 |
| Vietnam (1) | LMI | Asia |  |  |
| Thailand (2) | UMI | Asia |  |  |
| Peru (1) | UMI | South America | 1 study | 7.1 |
| Serbia (2) | UMI | Europe | 2 studies | 7.1 |
| **Study Designs** |  |  |  |  |
| Quantitative designs; Quantitative cross-sectional designs |  |  | 16 studies | 76.2 |
| Qualitative designs; Case Study, Interviews and FGDs |  |  | 3 studies | 14.3 |
| Mixed Methods |  |  | 2 studies | 9.5 |
| **Total** |  |  | **21 studies** | **100** |
| *LI- Low Income, LMI- Low Middle Income, UMI- Upper middle income. |  |  |  |  |

Models and frameworks utilised included the Competing Values Framework for management, Management Competency Assessment Project (MCAP), 360 Degree Competency Assessment, 2007 WHO framework for strengthening leadership and management capacity, World Management Survey (WMS) and Centres for Disease Control and Prevention (CDC) Managerial matrix of core competencies.

### Synthesis of results

#### Components of Individual and Institutional Capacity

In our findings, management capacity of PHC facilities consisted of individual and institutional capacity. The areas of individual and institutional capacity in the studies are illustrated in Table 2 and Table 3.

**Table 2:** Overview of the different components of individual capacity.

| Individual Capacity |
| --- |
| Communication. Financial management. Human resource management. Leadership. Planning, target setting and evaluation. Change management. Knowledge of the healthcare environment. Problem Solving. Professionalism. Community and client engagement. Stakeholder engagement. Performance management. Operations and administration. Decision making. Information management. Promotion and prevention of disease. Partnership and Collaboration. Job Motivation and Satisfaction. Resource management. Oversight and coordination. Situational analysis. Time management. Supportive management. Time management. Supportive management. Emotional intelligence. Proactive approach. Clinical services. Service provision. |

**Table 3:** Overview of the different components of institutional capacity.

| Institutional Capacity |
| --- |
| Access to adequate resources. Awareness of roles and responsibilities. Support to managers. |

Several components in individual and institutional capacity were similar and related to one another; these were grouped as illustrated in Fig 2.

**Fig 2:**
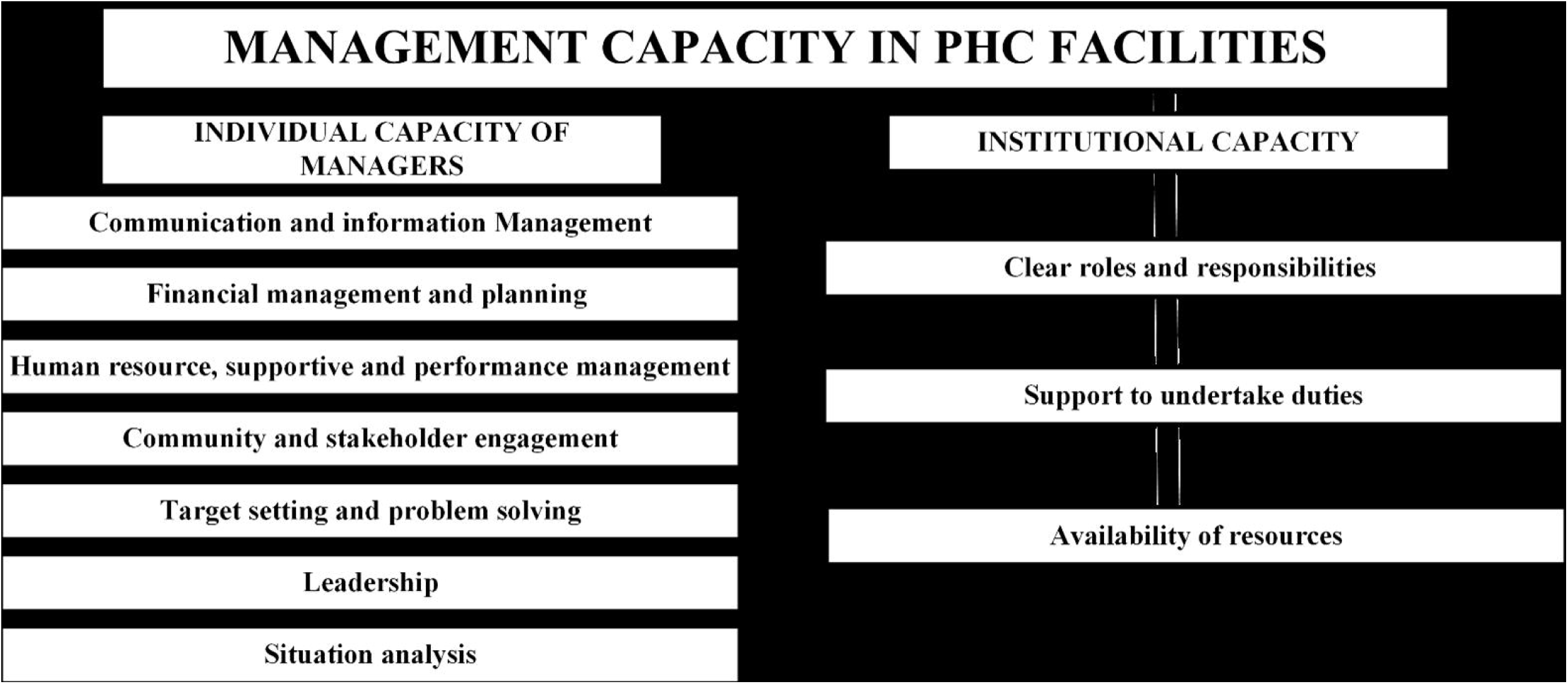
Components of Management Capacity of PHC Facilities.

##### a. Individual Capacity of Managers

Individual capacity primarily consisted of the managerial competencies of the facility managers. Managerial competency referred to the knowledge, skills, attitudes and behaviours that managers required to perform management tasks [10,30]. There were 32 competencies identified from the studies. The most included competency was human resource management which was in 14/21 (66.7%) of the studies [2,11,17,31–36]. These competencies were grouped into seven groups: communication, financial management and planning, human resource, supportive and performance management, community and stakeholder engagement, target setting and problem solving, leadership and situational analysis. These are discussed as follows.

###### i. Communication

Ability to effectively communicate with subordinates, the community, policymakers and supervisors was an important managerial competency [17,31–36]. Key communication skills included managers’ ability to listen attentively to different actors, share ideas, remain calm in emergencies, create open pathways of communication with actors, and provide accurate information [17,31–36]. Communication was especially important in convincing the public to take up public health services and sharing facility information with the community and other stakeholders [34].

However, despite its recognized importance, numerous studies revealed substantial deficiencies in managers’ communication skills, as assessed by themselves, their supervisors, and their subordinates [32,37]. Discrepancies were notable, with self-assessment ratings often being higher than evaluations from supervisors and subordinates [37]. Managerial experience was found to influence communication competency, with those having more experience generally rating themselves higher in this regard [17]. Overall, the findings underscore the pivotal role of communication in managerial effectiveness, influencing various aspects of organizational functioning, relationship-building, and public engagement.

###### ii. Financial management and planning

The ability of facility managers to effectively plan, budget and advocate for more financial resources for their facility was an important managerial competency [2,11,17,31–33,35,38]. Across the studies, necessary financial management skills included budget management, financial resources mobilization, reporting, prudent purchasing of needed facility drugs and equipment, management of financial risks and financial education for staff [2,11,17,31–33,35,38]. Budget management was to be done following the existing financial legislation [17,32]. These skills were important in enabling managers to develop realistic budgets, track and report, mobilize for more financial resources, manage financial risks and strategically purchase drugs and equipment based on facility needs and local priorities [11,19,33].

However, a good portion of managers lacked these financial management skills but those with significant managerial experience considering themselves more competent. For example, the studies by AwadAllah & Salem, 2021 and Adeniran et al., 2022, reported more than 50% of the managers were not competent in creating realistic budgets, financial reporting and mobilizing for more financial resources. The lack of budgeting and reporting competencies predisposes the facility to poor accountability, transparency and inefficiencies. Notably, managers who had more than five years managerial experience perceived themselves as more competent compared to those with little or no experience [17]. This was important as they could mentor other less experienced managers to help improve their competencies.

Planning as a competency also intersected with financial management, as managers had to consider the available annual facility budget while planning for the facility [17]. Managers had to balance between the available resources and the competing priorities in their planning [17,33,39].

###### iii. Human resource, supportive and performance management

Another important managerial competency was the manager’s ability to train, enhance performance, and foster a conducive work environment for human resources under their supervision [2,11,14,17,18,32,33,40]. In the studies, key human resource management (HRM) skills included performance monitoring, providing feedback, supportive supervision, staff in-service training, managing absenteeism, delegating, leading, and mentoring staff [2,11,14,17,18,32,33,40].

Managers possessing these skills implemented in-service training based on their staff skill needs, provided constructive feedback, resolved conflict, proactively managed absenteeism, and mentored staff, all of which contributed to improved staff performance [19]. For instance, in Lopes et al., 2020 and Foster et al., 2017, HRM competent managers conducted ongoing in-service training, provided staff with specified job descriptions, monitored and improved staff productivity and staff retention. Managers with more managerial experience and higher level positions were more competent [39]. However, many other managers lacked these competencies. For instance, AwadAllah & Salem, 2021, reported 60.1% of managers have limited human resource management skills similar to Munyewende et al., 2016, where more than half of supervisors and subordinates reported facility managers as not competent in these skills. When managers lack these competencies, staff are less motivated, perform below average and miss out on training opportunities to improve their skills. Thus, human resource management was a critical competency for managers, enabling them to create a conducive work environment for their staff and enhance staff performance.

###### iv. Community and stakeholder engagement

Another important managerial competency was the manager’s ability to involve the community and stakeholders in facility activities, gain their support, and feedback and implement changes according to the received feedback [11,14,18,33,38,40]. Across the studies, skills in community and stakeholder engagement included community outreaches, gaining community trust and satisfaction, getting community feedback and implementing changes, and establishing community health committees and advisory boards [11,14,18,33,38,40]. Competent managers ensured there were functioning community health committees, health advisory boards and clinic committees [14,18,38,41]. These committees supported the involvement of the community in decision-making, strengthening community linkages and improving service delivery [14,42]. Competent managers also collected feedback from the community using different tools such as suggestion boxes and implemented changes based on this received feedback, better addressing community health needs and priorities [18,41].

The performance of the managers in this competency was still mixed. Across the board, the managers scored poorly in ensuring community members and leaders were actively involved in facility activities including meetings, committees and advisory boards [14,18,33]. The managers however performed well in listening and responding to feedback [14,33]. Responding to this feedback is important as it meant care was responsive to the community’s needs and they were empowered to take an active role in the design of health services. Community engagement was therefore an important competency for the managers, but deficiencies were prevalent.

###### v. Target setting, problem-solving

Another managerial competency was the ability of managers to set facility goals, objectives, and targets, and implement timely corrective actions to address encountered risks and problems [14,17,33,34,38,43]. Key skills in this competency included setting of facility mission, vision, goals, action plans, improvement targets, and accountability to a specified population [17,33,34,38,43]. These goals addressed community needs and aligned with sub-district, district, and provincial targets and priorities. Problem-solving competency was also required in identifying the root causes of problems and taking timely corrective action [2,17,32].

There were notable deficiencies in the competency of managers across studies. For example, in the study by AwadAllah & Salem, 2021, 58.7% of managers reported having low managerial competency in planning and target-setting, and 56.6% reported low competency in problem-solving. In Munyewende et al., 2016, 54.1% of supervisors and 57.3% of subordinate nurses rated managers as not competent in problem-solving. This was a threat as such managers would not effectively implement corrective action to encountered problems including those affecting service delivery. Target setting and problem-solving skills were important individual capacities but deficiencies were prevalent.

###### vi. Leadership

Another important managerial competency was the manager’s ability to lead and motivate staff to achieve set missions and visions, consult, and handle difficult situations [17,31–34,43]. Key areas of leadership across the studies included motivating staff, achieving missions and visions, consultation in decision-making, feedback and coaching, delegation, and dealing with difficult workplace situations [17,31–34,43]. Competent managers set clear missions and values, consult staff when making strategic decisions, and empower talented staff [31,43]. Infection risks to patients were also reduced in facilities with managers competent in leadership [17,43]. The level of competency of managers across the studies was inconsistent, with some reporting high competency levels and others reporting low competency levels. For example, Kingu et al., 2023, found 75.48% of the managers to be competent in leadership while Munyewende et al., 2016, reported that more than half of the supervisors rated facility managers as not competent in leadership. Experience was also a key determinant, as more experienced managers self-rated higher in perceived competency levels. Leadership played a key role in motivating staff to achieve set goals; however, some managers lacked proficiency in this area.

###### vii. Situational analysis

Also, managers’ situational awareness of the healthcare environment, its influence on the functioning of facilities, and how to implement change were important managerial competencies [2,31,32]. Across the studies, skills in situational analysis included the manager’s knowledge of the regulatory and administrative environment, the role of different healthcare professionals and healthcare activities related to their work area. It also included planning, implementing, and monitoring change [2,31,32]. Good awareness meant a better understanding of national guidelines, strategies, and policies, and how they relate to the manager’s work area. Facility managers competent in change management effectively recognized the need for change, developed, implemented, and monitored change strategies, and also managed resistance to change [31]. The competency of managers across the studies was mixed. Lopes et al., 2019, reported poor competency in knowledge of the healthcare environment, while Kingu et al., 2023, found that more than 65.68% were competent. Despite the good competency of some managers, much should be done to improve the competency of those with lower levels.

There was an interrelationship between the different competencies we found. The various competencies complemented each other, making it essential for managers to possess most of them to effectively oversee the operations of PHC facilities. For example, communication as a competency was also important in human resource management and community engagement as managers needed to effectively communicate with staff and the community.

##### b. Institutional capacity

Institutional capacity included the availability of management support and resources at the facilities for managers to carry out planned activities and their awareness of their roles and responsibilities. Across the studies, facility, sub-district, district and regional management teams were responsible for the provision of resources to PHC facilities [2,15,44]. They were also responsible for providing functional management support to PHC facility managers depending on the existing type of decentralization and the extent of their autonomy [2,11,45]. Functional management support systems include the provision of funds, logistical support and infrastructure that managers require to carry out planned facility activities, clear roles and responsibilities and management support [2,11,40,45].

###### i) Clear roles and responsibilities

Ensuring that managers are aware of their roles, responsibilities and degree of authority was another important aspect of functional support systems. Managers were expected to be conversant with their roles and responsibilities as these guided them in their work (Adeniran et al., 2022; Foster AA et al., 2017; Heerdegen ACS et al., 2020; Liu L, Desai MM, Fetene N, et al., 2022). They also needed to be aware of the extent of their authority and accountable. These roles, responsibilities and degree of authority are to be contained in the job descriptions. In the studies, this awareness and availability of job descriptions were varied. Adeniran et al., 2022 and Heerdegen et al., 2020 reported that majority of managers were provided with clear job descriptions. This was opposite to the findings of Foster et al., 2017 where managers lacked an explicit job description which augmented the difficulties they faced as they transitioned from clinical to management roles. However, even with job descriptions, managers were still tasked with additional responsibilities not within their scope (Adeniran et al., 2022). moderate/large extent. This is a concern as these managers may not have been adequately prepared and supported to carry out these additional duties. Therefore, the availability of clearly defined job descriptions is a requirement as part of institutional capacity.

###### ii) Availability of resources

Availability of the required resources to carry out planned facility activities was an important part of functional support systems as provided by the managing authorities [2,11]. Studies assessed whether the PHC facilities had adequate funds, logistics and infrastructure necessary for managers to carry out facility activities. For public facilities, the sub-national and national management structures were often responsible for providing these resources [2,33,44]. As integral inputs, the availability of these resources contributed to what extent planned facility activities were implemented. However, the managing authorities had not readily provided these required resources to the facilities thereby limiting the managers’ running of the facilities and service delivery. This led to challenges such as shortage of essential drugs and equipment reducing the availability, accessibility and quality of services. Inadequate and irregular financial resources was most reported across studies. In Adeniran et al., 2022, 71.8% of facility managers reported that they occasionally had adequate funds to carry out planned activities. This irregular disbursement from the national governments is also documented by Altobelli, 2024 who add that it delayed the purchasing of essential drug, supplies and equipment. The limited availability of adequate funds, logistics, and infrastructure was therefore an impediment to facility operations.

###### iii) Support to undertake duties

Supporting managers when they require assistance to complete their assignments was another important aspect of functional support systems [2,11]. Managers were to be supported in assignments such as human resource management, data management, procurement of drugs and other commodities, community involvement, planning, and budgeting [2,11]. This support also included supportive supervision, feedback, and mentoring from their supervisors in the higher levels of management at the sub-district and district levels [2,11,40]. Such support was important in collective problem solving and resolving issues that previously needed escalation. However, the level of this support varied across the studies. In Adeniran et al., 2022, 60.6% of managers reported occasionally receiving support in conducting planning and budgeting. Conversely, supportive supervision, feedback, and mentoring were better provided across the studies, with 89.7% of managers in Heerdegen et al., 2020 indicating ‘to a moderate/large extent,’ and 63.3% of managers in Adeniran et al., 2022 reporting ‘always.’ Thus, managers were better supported in some areas while other areas received little to no support. Nonetheless, considering that providing optimal support can create a conducive work environment for managers, it was important for managing authorities to make this available.

In summary, these three areas of institutional capacity were interrelated and interdependent. Improving one required a concurrent improvement in the others for better capacity. For instance, managers needed to not only understand their roles and responsibilities but also have access to the resources essential to effectively fulfil those duties. Similarly, they also needed management support to effectively undertake these duties.

Management capacity of PHC facilities required both individual and institutional capacities to be in place. Managerial competencies were essential for the daily operation of these facilities and the efficient use of available resources. Institutional capacity determined the availability of these resources for example via district and sub-district management teams in public PHC facilities [2,10,11,44]. Functional support systems were only useful if the needed management competencies were present. Considering this interrelatedness, strengthening either individual or institutional capacity without the other was not likely to work [2,10].

## DISCUSSION

The objective of our review was to synthesize evidence on the components of management capacity of PHC facilities in LMICs. We have reviewed studies that reported various components of management capacity of PHC facilities and categorised these into individual and institutional capacity. Individual capacity consists of the competencies of managers such as communication, human resources management, financial management and leadership. Institutional capacity on the other hand includes the support and resources provided to managers. The existing individual and institutional capacities were both inadequate. They also affected the availability of medicine and equipment, perceived quality of care and satisfaction and other service delivery elements.

The components we have identified in our review are similar to the dimensions of the WHO leadership and management framework. The WHO leadership and management framework is a recognised framework that outlines four dimensions essential for the improvement of management capacity of health systems in LMICs[10]. These dimensions are; appropriate competencies, functional support systems, an enabling work environment and an adequate number of managers. In our findings, individual capacity mainly consisted of managerial competencies which is a dimension in the framework. Also, our findings on institutional capacity capture both the functional support systems and supportive work environment dimensions. However, the dimension on adequate number of managers was not extensively covered in the studies we reviewed. This is plausibly a gap that future research can explore.

Our findings better identify individual and institutional capacities as part of management capacity compared to other reviews that have focused solely on managerial competencies. Globally, existing reviews have been conducted to summarise evidence on managerial competencies but less of institutional capacities. A systematic review and best-fit framework synthesis by Kakemam et al., 2020 presented seven core competencies that were similar to the competencies we identified in our study [20] professionalism [20]. These competencies influence service delivery which is an important element of facility performance. While their findings are important in detailing the required skills and capabilities, they don’t report on institutional capacities. The same is seen in the scoping review by Ndayishimiye et al., 2023 who categorize competencies as ‘soft’ and ‘hard’ competencies but doesn’t explore the role of institutions in supporting managers and providing the needed resources [21]. Promisingly, our review has provided a more robust discussion of what these institutional capacities are and how they contribute to management capacity of PHC facilities in LMICs. Institutional capacities are an important part of management capacity and should be considered both in research and program implementation.

Management capacity gaps in the studies are concerning considering the negative effects it will have on service delivery. Existing evidence has shown that better management capacity is an important ingredient for improved health services [10]. Facilities with good management capacity have better coverage and quality of services, patient respect and indiscrimination, and better patient outcomes [11,12,46]. The existing management capacity gaps found in our review are therefore a threat to the coverage, quality and patient-centeredness of the services provided by the facilities. This limitation in service delivery such as inaccessible, unavailable, unsatisfying and low-quality care will hamper the achievement of UHC where PHC is an important vehicle. Therefore, these gaps should be improved upon to better services at the facilities.

Furthermore, gaps in the financial management competencies of managers and lack of institutional support in planning and budgeting across the studies will likely reduce the benefits gotten from any implemented financial autonomy reforms. Financial autonomy provides facilities with more influence and control over the mobilization, allocation and spending of financial resources [47–49]. Evidence has shown that in some cases, financial autonomy may contribute to technical efficiency and responsiveness [50,51]. However, these benefits are better realised where the facility management is competent and accountable [52]. The lack of competent managers may lead to unnecessary wastage of these financial resources and inefficiency [6,10]. As such, existing gaps in the financial management competencies of the managers and the limited institutional functional support systems should be addressed for countries planning or implementing autonomy reforms.

The strength of our review includes the systematic and rigorous process we undertook, and the novelty of our results. Using the Arksey and O’Malley framework, we carried out the review in five systematic steps, ensuring rigour and replicability [22]. We reported our results based on the PRISMA-ScR reporting guidelines, ensuring transparent and comprehensive reporting [25]. Also, our review is the first one to summarize the extent of literature on management capacity of PHC facilities in LMICs, its components and performance across these components. It provides useful information to policymakers on evidence-based gaps that can be acted upon and also lays the foundation for further research.

Nevertheless, these findings should be considered in the context of the following limitation. There is a possibility that we may have missed important empirical literature on management capacity in LMICs. To mitigate this, we conducted an extensive search and hand-checking of reference lists for any papers that may have been missed.

There is a need for more research on management capacity of PHC facilities and interventions to improve existing gaps. From our review, the literature on management capacity of PHC facilities in LMICs is still limited, warranting for more studies considering its important role in PHC and UHC. More studies are also needed to understand better the relationship between the financial management competencies of managers and the accrued benefits of facility financing autonomy reforms. Improvement interventions are needed to improve the found management capacity gaps. Interventions such as management training and mentorship of new managers by experienced managers have shown the potential to improve the managerial competencies of the managers.

## CONCLUSION

From our review, management capacity of PHC facilities in LMICs consists of individual and institutional capacity. Individual capacity includes the competencies of the managers such as communication, human resource management, financial management and community engagement. Institutional capacity includes functional support systems such as making resources available to managers and offering logistical support. Regarding performance, managers performed poorly in several competencies such as communication, financial management, human resource management and community and stakeholder engagement. Their institutions also failed to effectively provide them with the resources and logistical support they needed. The scarcity of these resources reduced the availability, accessibility and quality of services. There is still a scarcity in the number of studies and more research to be conducted. Furthermore, interventions such as training and supportive supervision should be considered in improving managerial competencies of managers.

## Supporting information

S1 File- PRISMA Checklist

S2 File- Data Extraction

S3 File- Search terms for PubMed

S4 File- Search terms for Web of Science

S5 File- Search terms for Scopus

S6 File- Search terms for Google Scholar

## AUTHOR CONTRIBUTIONS

**Conceptualization:** Harrison Ochieng, Anita Musiega, Jacinta Nzinga, Benjamin Tsofa, Edwine Barasa

**Data curation:** Harrison Ochieng, Anita Musiega

**Formal Analysis** : Harrison Ochieng, Anita Musiega

**Methodology:** Harrison Ochieng, Anita Musiega, Jacinta Nzinga, Benjamin Tsofa, Edwine Barasa

**Visualization:** Harrison Ochieng

**Writing-original draft preparation:** Harrison Ochieng

**Writing-review & editing:** Harrison Ochieng, Anita Musiega, Jacinta Nzinga, Benjamin Tsofa, Edwine Barasa

**Supervision:** Anita Musiega, Jacinta Nzinga, Benjamin Tsofa, Edwine Barasa

## DATA AVAILABILITY STATEMENT

The data that support the findings of this study are available on request from the corresponding author.

## ETHICS STATEMENT

This being a secondary synthesis, no ethical approval was required.

## SUPPLEMENTARY INFORMATI ON

**S1 File- PRISMA Checklist**

**S2 File- Data extraction form and study characteristics**

**S3 File- Search terms for PubMed**

**S4 File- Search terms for Web of Science**

**S5 File- Search Terms for Scopus**

**S6 File- Search Terms for Google Scholar**

