## Supplementary material for "Management capacity of healthcare facilities in Low- and Middle-Income Countries; A scoping review": S2 File- Data Extraction

**Supplemental File 1: Data extraction form and summary of study characteristics**

| **Author and Year** | **Country and World Bank Classification** | **Study Design** | **Participants** | **Theories, Models and Frameworks** | **Capacities** |
| --- | --- | --- | --- | --- | --- |
| AwadAllah & Salem, 2021 | Egypt (Lower Middle Income) | Cross Sectional Study | Primary Healthcare Facility Managers |  | **Individual Capacity;**   i. Communication- Listening to concerns, sharing ideas, overcoming communication barriers and building relationships. 56.5% had high competency. 60.9% considered it important. ii. Financial management- Budgeting in accordance with financial legislation, realistic projections, evaluating expenditure and staff financial education. 54.3% had low competency. 41.3% considered it important. iii. Human resource management- managing absenteeism, delegation, feedback, performance monitoring and in-service training. 60.1% had low competency. 57% considered it important. iv. Leadership- visionary, consulting, motivating staff, dealing with difficult patients and reducing infection risk. 50% had low competency. 41.3% considered it important. v. Planning and priority setting- Information management, task prioritization, community needs identification, set mission, vision, goals and action plans. 58.7% had low competency. 37.0% considered it important. vi. Problem solving- risk monitoring, manage emergencies and correction actions. 56.6% had low competency. 60.9% considered it important.   Across all competencies, experienced managers rated highly on self-rating and perceived importance of the respective competency. |
| Lopes et al., 2019 | Timor-Leste (Low Income Country) | Cross Sectional Survey | Primary Healthcare managers |  | **Individual Capacity**   i. Communication- business communications, listening, communicating clearly with stakeholders. Most respondents rated as competent ii. Financial management- budgeting in accordance with financial legislation and registering assets. Majority rated intermediate competency (not competent) iii. Human resource management- in-service training, supervision, performance management and availing job descriptions. Majority rated as competent. iv. Leadership- adherence to legal and regulatory standards, encouraging commitment, consulting, constructive feedback, mentoring talent and dealing with difficult situations. Majority rated intermediate competency (not competent). v. Knowledge of the organization- regulatory and administrative environment, socioeconomic environment, health systems, community health standards and delivery, role of different cadres and competition. Majority rated intermediate competency (not competent). vi. Professionalism- conducting responsibilities, professional development, conflict management as per laws, mentoring and contributing to professional knowledge. Majority rated intermediate (not competent). |
| Mabuchi et al., 2020 | Nigeria (Lower Middle-Income Country) | Cross Sectional Design | Primary Healthcare Centers Managers |  | **Individual Capacity**   i. Communication- Atmosphere and process for open communication. Average score was 2.23 in 1-3 scale. ii. Financial management- Utilizing PBF funds according to guidelines and updating financial records. PBF guidelines dictated proportions to be spent on drugs and supplies and bonuses. Average score was 2.27. iii. Human resource management- staff involvement in PBF decisions and teamwork building. Average score was 2.31 iv. Planning and target setting- business plans and target updates. Average score was 2.22 v. Community/client engagement- community outreach, community trust and satisfaction, client recruitment and retention. Average score was 1.96 vi. Stakeholder engagement- Engaging supervisor and community leaders. Average score was 1.87. vii. Performance management- Performance tracking and performance review. Average score- 2.18 |
| Kingu et al., 2023 | Tanzania (Lower Middle-Income Country) | Cross Sectional Descriptive research design | Primary Healthcare Facility Managers | Management Competency Assessment Project (MCAP) Framework | **Individual Capacity**   i) Communication- active listening, trust and rapport, negotiation and conflict resolution. 74.5% rated as competent ii) Leadership- Setting clear goals and objectives, inspiring and motivating staff, constructive feedback, coaching development, creating supportive work environment and delegation. 75.48% rated as competent. iii) Change management- recognizing need for change, involving stakeholders, implementing and managing change resistance. 74.55% rated competent iv) Decision making- Informed decisions from collected data and evidence 67.65% rated as competent v) Resource management- planning, budgeting and allocation of resources, and resolving operational issues. 62.75% of managers rated as competent vi) Knowledge of the healthcare environment- professionalism, ethical issues, planning and service provider management. 65.68% rated as competent. |
| Mohd-Shamsudin & Chuttipattana, 2012 | Thailand (Upper Middle-Income Country) | Cross Sectional Study | Primary Care Managers |  | **Individual Capacity**   i. Communication- increasing public`s knowledge and support for public health issues. ii. Leadership- facilitate staff understanding and commitment to agency goals and mission. iii. Planning- supporting assessment, planning and evaluation process and supports colleagues doing the same. Refining goals/objectives based on evaluation results iv. Information management- confidentiality of client data and utilization to aid client`s lifestyle choices v. Partnership and collaboration-cooperates with other organizations and other partners to aid coordinating goals and services. |
| Adeniran et al., 2022 | Nigeria (Lower Middle-Income Country) | Descriptive Cross-Sectional Design | Primary Healthcare Facility managers |  | **Individual Capacity**   i) Financial management- budgeting, mobilizing financial resources and financial reporting. Average score was 12.9(4.7)- max of 30 ii) Human resource management- performance management, motivating staff, availing job descriptions, assigning supervisors and annual appraisal. Average score was 45.2(11.2)- max of 78. iii) Problem solving- Identifying root causes of problems and developing clear problem statements and solutions. Average score was 23.6(5.4)- max of 36. iv) Information management- Gathering, analyzing and reporting facility information to stakeholders and communities to improve health services. Average score was 12.9(4.7)- max of 22. v) Job motivation and satisfaction- Motivating to work, satisfaction and met expectations. Average score 26.7(3.1), max of 60. vi) Oversight and coordination- addressing gaps, feedback to stakeholders and coordinating meetings and workshops. Average score 5.0(3.0)- max of 28 vii) Situational analysis- Understanding national guidelines and health priorities. Average score 15.0(4.3)- max of 24   **Institutional Capacity**   i) Access to adequate resources- finances, logistics and infrastructure ii) Awareness of roles and responsibilities- available job descriptions and national guidelines, and additional roles. iii) Supporting managers when they require assistance- support in planning and budgeting, procurement, human resource management, community involvement. |
| Mubiri et al., 2024 | Ghana (Lower Middle Income) | Cross Sectional Study | Primary Healthcare Facility Managers |  | **Individual Capacity**   i) Resource management- maintenance, repair and replacing of equipment. Avoiding drug and supplies stockouts. ii) Time management- Managing shifts ensuring breaks, regular leave from work and managing frequency of on-call nights. iii) Supportive management- Dealing problematic personnel, supporting staff understand and implement clinical guidelines, treating staff openly and fairly, resolving staff-management conflict and staff in-service training. |
| Daire & Gilson, 2014 | South Africa (Upper Middle Income) | Qualitative Case Study Approach | Primary Healthcare Facility Managers | Leadership and organizational theory | **Individual Capacity**   i) Financial management- Awareness of facility resource needs, prioritize needs, meetings to track facility budget and controlling expenditure. ii) Human resource management- proactively managing absenteeism, interpersonal relationships, role modelling, coaching, supervision, motivating and identifying staff development needs. iii) Planning- orienting service delivery based on community health needs identified from community profiles, district and sub-district priorities, participating in annual planning at the district and subdistrict levels. iv) Community engagement- facilitating community engagement and community health committees, collecting feedback from clients, addressing community and client complaints. v) Resource management- Ensuring drugs and equipment are available and functioning. Procurement and inventory for drugs and supplies and the presence of a pharmacist or pharmacy assistant in the facility. vi) Service provision- Knowledge on target population and their health needs, orienting service delivery to the population, monitoring service targets and submitting reports to sub-district office and assessing performance of facility. |
| Kitreerawutiwong et al., 2015 | Thailand (Upper Middle Income) | Mixed Methods | Primary Healthcare Facility Managers, policy makers, academicians and sub-district administrators |  | **Individual Capacity**   i) Communication- clear information to patients and public, garnering facility support from the public, communicating public health issues to policy makers and developing trust with different stakeholders. ii) Financial planning- Cost-effective programs, gaining financial support from facility committee and managing financial risks. iii) Leadership- Clarifying mission, vision and goal, understanding and communicating policy to staff, innovate, coaching and performance assessment of healthcare workers. iv) Organizational development and Professionalism- courtesy, confidentiality of clients, respect, protect client`s right and work and achieving reliable performance. v) Information management- Ensuring fast and convenient access to information, applying information technology to serve health needs and using community media to community health issues and community health information management system. vi) Emotional intelligence- Listen to other`s ideas and accepting results of performance evaluation |
| Munyewende et al., 2016 | South Africa (Upper Middle Income) | Cross Sectional Study | Primary Healthcare Clinic Nursing Managers | **1.** The 2007 World Health Organization (WHO) conceptual framework for strengthening leadership and management capacity. **2.** 360 Degree competency Evaluation | **Individual Capacity**   i) Human resource management- staff in-service training, performance management, manage absenteeism, delegation and constructive feedback. 26.7% of managers rated themselves as not competent. 59.2% of supervisors and 56.3% of subordinates rated managers as not competent. ii) Leadership- Implementing health improvement vision, consulting, dealing with difficult situations and reducing infection risk. 29.5% of managers rated themselves as not competent. 53.1% of supervisors and 49.5% of subordinates rated managers as not competent. iii) Planning and priority setting- Involving stakeholders in identifying needs and priorities, defending clinic interests, meeting agreed tasks and priorities. 31.1% of managers rated themselves as not competent. 62.2% of supervisors and 51.5% of subordinates rated managers as not competent. iv) Problem Solving- monitoring safety issues, calmly managing emergencies, identifying root causes, timely implementation of corrective actions and conflict resolution. 28.6% of managers rated themselves as not competent. 54.1% of supervisors and 57.3% of subordinates rated managers as not competent. |
| Heerdegen et al., 2020 | Ghana (Lower Middle Income) | Cross Sectional Study | Primary Healthcare District managers |  | **Individual Capacity**   i) Communication- Belongingness, cooperation and frequency of communication with team members. Average score was 4.2(0.6) out of 5. ii) Financial management- Developing and managing targets, mobilizing financial resources and financial reporting. The average score was 3.9(1.0) out of 5. iii) Human resource management- identifying human resource needs, regular off for staff to avoid fatigue, lobbying and posting human resource and monitoring personnel performance. The average score was 4.2(0.7). iv) Service delivery and community involvement- Involvement of community in decisions and gaining feedback from community and ensuring staff access to guidelines. Average score was 4.4(0.6). v) Information management- gathering, analysing and reporting facilities information to stakeholders. Average score was 4.5(0.7). vi) Resource management- Ensuring availability of drugs and supplies, repair and maintenance of equipment. Average score was 4.1(0.8).   **Institutional Capacity**   i) Access to adequate resources- finances, logistics and infrastructure ii) Awareness of roles and responsibilities- available job descriptions and national guidelines, and additional roles. iii) Supporting managers when they require assistance- support in planning and budgeting, procurement, human resource management, community involvement. |
| Macarayan et al., 2019 | Ghana (Lower Middle Income), Uganda (Low Income) and Malawi (Low Middle Income) | Analytical Cross Sectional Study | Facility heads and women who sought care | World Management Survey (WMS) | **Individual Capacity**   i) Human resource management- in-service training, performance management and supportive supervision for staff. Average score was 0.89(0.17) in a 0-1 scale. ii) Target setting- Annual budget and accountable for specific population. Average score was 0.74(0.25) iii) Community engagement- collecting client opinion, sharing performance information, community advisory boards and inclusion of community member in staff meetings. Average score was 0.65(0.20). iv) Operations- handwashing area, facility open every day, manager trained on management and time spent on managerial activities. Average score was 0.73(0.18). v) Monitoring- tracks revenue and expenditure, quality improvement, auditing, reporting disease outbreaks and monitoring facility performance. Average score was 0.41(1.21). |
| Foster et al., 2017 | Zambia (Low Income Country) | Qualitative; Focus Group Discussions and Interviews | Rural facility heads, ministry officials, provincial and district administrators |  | **Individual Capacity**   i) Human resource management- delegating, mentoring, motivating and retaining. ii) Leadership- Responding to community needs, engaging community in decision making and leveraging facility head position to influence change in the community. iii) Resource management- Managing flow of medicines and commodities, and facility repairs. |
| Tetui et al., 2017 | Uganda (Low Income Country) | Qualitative: Observations and interviews | Primary healthcare managers | The Competing Values Framework for Management | **Individual Capacity**   i) Collaborate function- Enhancing commitment to project goals and team cohesion. ii) Control function- Planning, budgeting, coordination and implementation of activities iii) Compete function- Defining and attaining set goals and confidence in decision making iv) Create function- Unmask existing resources for health needs and becoming more adaptable to client needs. |
| Liu et al., 2017 | Ethiopia (Low Income Country) | Cross Over design with Control group | Managers from Health centers and Woredas |  | **Individual Capacity**   i) Human resource- Availed job descriptions, performance evaluation, staff uniforms and motivation package for staff according to Ethiopia Health Center Reform Implementation Guidelines (EHCRIG) guidelines. Before intervention, score was 30% to 40% standards met which improved to 60% to 70% post intervention. ii) Leadership- Oversight, regular meetings, financial transparency including visible user fees and presentation of procurement plans to governing body. 40% to 50% standards met before intervention and 65% to 75% post intervention. iii) Community engagement- Linking health posts to heath extension workers and professional support for Primary Health Care Units. 45% to 55% before intervention and 70% to 80% post intervention. iv) Performance management- performance monitoring and quality improvement using set indicators and client satisfaction surveys. 40% to 50% standards met pre-intervention and 60% to 70% post-intervention. v) Resource management- inventories, maintenance and repair of equipment. Equipment operators well trained and available. Availability of water and electricity. Preintervention 10% to 20% and 30% to 40% post-intervention. vi) Clinical, laboratory and pharmaceutical services- Standard operating procedures, guidelines, resources and staffing. 40% to 50% standards met post intervention up from 30% to 40% pre-intervention. |
| Uy et al., 2019 | Ghana (Lower Middle-Income Country) | Quantitative Psychometric validation study | PHC facilities and their heads |  | **Individual Capacity**   i) Human resources management- staff training, supervision, staff performance management and staff ability to complete assignments. Score was 0.89 in 2016 and 0.93 in 2017 in a 0-1 scale. ii) Target setting- monitoring key coverage indicators, annual budget for running costs, accountability for specified population, improvement targets for service delivery and shared with staff. Score was 0.74 in 2016 and 0.77 in 2017. iii) Community engagement- Collecting client opinions, changes based on client opinions, sharing performance information with community, community advisory board and community member attending staff meetings. Score of 0.65 in 2016 and 0.65 in 2017.   iv) Operations- handwashing area, facility open every day, manager trained on management and time spent on managerial activities. Score was 0.73 in 2016 and 0.71 in 2017.  v) Monitoring- tracks revenue and expenditure, quality improvement, auditing, reporting disease outbreaks and monitoring facility performance. Score was 0.81 in 2016 and 0.79 in 2017. |
| Kim et al., 2022 | Uganda (Low Income Country) | Cross Sectional Survey | Health Facilities and managers |  | **Individual Capacity**  i) Human resources management- staff training, supervision, staff performance management and staff ability to complete assignments. Median score was 0.833 in a 0-1 scale.  ii) Target setting- monitoring key coverage indicators, annual budget for running costs, accountability for specified population, improvement targets for service delivery and shared with staff. Median score was 0.667.  iii) Community engagement- Collecting client opinions, changes based on client opinions, sharing performance information with community, community advisory board and community member attending staff meetings. Score of 0.700  iv) Operations- handwashing area, facility open every day, manager trained on management and time spent on managerial activities. Median score was 0.50  v) Monitors and tracks revenue and expenditure, quality improvement, auditing, reporting disease outbreaks and monitoring facility performance. Median score was 0.83 |
| Dikic et al., 2019 | Serbia (Upper Middle Income) | Cross Sectional Design | Primary Care managers |  | **Individual Capacity**  i) Communication- Oral presentation, interview or writing articles to media, planning and implementing communication strategy. Perceived competency score was 4.03(0.76) in a 0-5 scale.  ii) Leading- Aligning teamwork, motivating employees and sharing vision and mission. Perceived competency was 4.22(0.61) in a 0-5 scale.  iii) Planning and priority setting- setting priorities, establishing workplans, applying decision analysis techniques, designing programs, and strategic, business and workplans. Perceived competency score was 1.03(0.66).  iv) Problem Solving- Identifying problems, root causes and factors and implementing corrective actions. Perceived competency score was 4.20 (0.61).  v) Performance assessment- evaluating employee performance, program and budget expenditures. Perceived competency score was 4.0(0.65). |
