## Supplementary material for "Management capacity of healthcare facilities in Low- and Middle-Income Countries; A scoping review": S3 File- Search terms for PubMed

**PUBMED Search Strategy**

| **#** | **Query** | **Date Searched** | **Results** |
| --- | --- | --- | --- |
| **S1** | ((("manager" OR "administrator" OR "supervisor" OR "coordinator" OR "head" OR "lead" OR "overseer" OR "director" OR "controller" OR "executive" OR "manag*" OR "administrat*" OR "supervis*" OR "coordinat*" OR "overs*" OR "direct*" OR "control*" OR "execut*" OR "operat*" OR "operat*" OR "steer*" OR "running" OR "chair*" OR "officer" OR "in-charge" OR "coordinat*" OR "chief" OR "superintendent")) AND ((("managerial capacity"[tiab:~2] OR "leadership capacity"[tiab:~2] OR "administrative capacity"[tiab:~2] OR "governance capacity"[tiab:~2]) OR ("manag*" AND "Capacit*") OR ("lead*" AND "capacit*") OR ("govern*" AND "capacit*") OR ("administrat*" AND "capacit*")) OR ("organi*" AND "capacit*") OR "capacity building"[MeSH Terms] OR ("manag*" AND "competenc*") OR ("facility manag*"))) | 24/10/2024 | 242,040 |
| **S2** | (("primary health care"[tiab] OR "primary care"[tiab] OR "primary healthcare"[tiab] OR "Primary health" OR "community health" OR "community health care"[tiab] OR "community healthcare"[tiab] OR "community care" OR "Community health cent*"[tiab] OR "community health clinics"[tiab] OR "health cent*"[tiab] OR "health cent*"[tiab] OR "health clinics"[tiab] OR "district health service*"[tiab] OR "district health" OR "preventive health service*"[tiab] OR "primary level" OR "primary-level" OR "frontline health" OR "frontline healthcare" OR "rural health*" OR "general practice" OR "family practice" OR "family medicine") AND (("Primary Health Care"[MeSH Terms]) OR ("Community Health Service*"[MeSH Terms]))) | 24/10/2024 | 183,508 |
| **S3** | ((Afghanistan[TIAB] OR Albania[TIAB] OR Algeria[TIAB] OR American Samoa[TIAB] OR Angola[TIAB] OR Antigua[TIAB] OR Barbuda[TIAB] OR Argentina[TIAB] OR Armenia[TIAB] OR Azerbaijan[TIAB] OR Bangladesh[TIAB] OR Belarus[TIAB] OR Byelarus[TIAB] OR Byelorussia[TIAB] OR Belorussia[TIAB] OR Belize[TIAB] OR Benin[TIAB] OR Bhutan[TIAB] OR Bolivia[TIAB] OR Bosnia[TIAB] OR Herzegovina[TIAB] OR Hercegovina[TIAB] OR BosniaHerzegovina[TIAB] OR Bosnia-Hercegovina[TIAB] OR Botswana[TIAB] OR Brazil[TIAB] OR Brasil[TIAB] OR Bulgaria[TIAB] OR Burkina[TIAB] OR Upper Volta[TIAB] OR Burundi[TIAB] OR Urundi[TIAB] OR Cambodia[TIAB] OR Republic of Kampuchea[TIAB] OR Cameroon[TIAB] OR Cameroons[TIAB] OR Cape Verde[TIAB] OR Central African Republic[TIAB] OR Chad[TIAB] OR Chile[TIAB] OR China[TIAB] OR Colombia[TIAB] OR Comoros[TIAB] OR Comoro Islands[TIAB] OR Comores[TIAB] OR Congo[TIAB] OR DRC[TIAB] OR Zaire[TIAB] OR Costa Rica[TIAB] OR Cote d'Ivoire[TIAB] OR Ivory Coast[TIAB] OR Cuba[TIAB] OR Djibouti[TIAB] OR Obock[TIAB] OR French Somaliland[TIAB] OR Dominica[TIAB] OR Dominican Republic[TIAB] OR Ecuador[TIAB] OR Egypt[TIAB] OR United Arab Republic[TIAB] OR El Salvador[TIAB] OR Eritrea[TIAB] OR Ethiopia[TIAB] OR Fiji[TIAB] OR Gabon[TIAB] OR Gabonese Republic[TIAB] OR Gambia[TIAB] OR Georgia[TIAB] OR Ghana[TIAB] OR Gold Coast[TIAB] OR Grenada[TIAB] OR Guatemala[TIAB] OR Guinea[TIAB] OR GuineaBissau[TIAB] OR Guiana[TIAB] OR Guyana[TIAB] OR Haiti[TIAB] OR Honduras[TIAB] OR India[TIAB] OR Indonesia[TIAB] OR Iran[TIAB] OR Iraq[TIAB] OR Jamaica[TIAB] OR Jordan[TIAB] OR Kazakhstan[TIAB] OR Kenya[TIAB] OR Kiribati[TIAB] OR Republic of Korea[TIAB] OR North Korea[TIAB] OR DPRK[TIAB] OR Kosovo[TIAB] OR Kyrgyzstan[TIAB] OR Kirghizstan[TIAB] OR Kirgizstan[TIAB] OR Kirghizia[TIAB] OR Kirgizia[TIAB] OR Kyrgyz[TIAB] OR Kirghiz[TIAB] OR Kyrgyz Republic[TIAB] OR Lao[TIAB] OR Laos[TIAB] OR Latvia[TIAB] OR Lebanon[TIAB] OR Lesotho[TIAB] OR Basutoland[TIAB] OR Liberia[TIAB] OR Libya[TIAB] OR Lithuania[TIAB] OR Macedonia[TIAB] OR Madagascar[TIAB] OR Malagasy Republic[TIAB] OR Malawi[TIAB] OR Nyasaland[TIAB] OR Malaysia[TIAB] OR Malaya[TIAB] OR Malay[TIAB] OR Maldives[TIAB] OR Mali[TIAB] OR Marshall Islands[TIAB] OR Mauritania[TIAB] OR Mauritius[TIAB] OR Mayotte[TIAB] OR Mexico[TIAB] OR Micronesia[TIAB] OR Moldova[TIAB] OR Moldovia[TIAB] OR Mongolia[TIAB] OR Montenegro[TIAB] OR Morocco[TIAB] OR Mozambique[TIAB] OR Myanmar[TIAB] OR Burma[TIAB] OR Namibia[TIAB] OR Nepal[TIAB] OR Nicaragua[TIAB] OR Niger[TIAB] OR Nigeria[TIAB] OR Pakistan[TIAB] OR Palau[TIAB] OR Palestine[TIAB] OR Panama[TIAB] OR Papua New Guinea[TIAB] OR Paraguay[TIAB] OR Peru[TIAB] OR Philippines[TIAB] OR Romania[TIAB] OR Rumania[TIAB] OR Roumania[TIAB] OR Russia[TIAB] OR Russian Federation[TIAB] OR USSR[TIAB] OR Soviet Union[TIAB] OR Union of Soviet Socialist Republics[TIAB] OR Rwanda[TIAB] OR RuandaUrundi[TIAB] OR Samoa[TIAB] OR Samoan Islands[TIAB] OR Sao Tome[TIAB] OR Principe[TIAB] OR Senegal[TIAB] OR Serbia[TIAB] OR Montenegro[TIAB] OR Yugoslavia[TIAB] OR Seychelles[TIAB] OR Sierra Leone[TIAB] OR Solomon Islands[TIAB] OR Somalia[TIAB] OR South Africa[TIAB] OR Sri Lanka[TIAB] OR Ceylon[TIAB] OR Saint Kitts[TIAB] OR St Kitts[TIAB] OR Saint Christopher Island[TIAB] OR Nevis[TIAB] OR Saint Lucia[TIAB] OR St Lucia[TIAB] OR Saint Vincent[TIAB] OR St Vincent[TIAB] OR Grenadines[TIAB] OR Sudan[TIAB] OR Suriname[TIAB] OR Surinam[TIAB] OR Swaziland[TIAB] OR Syria[TIAB] OR Syrian Arab Republic[TIAB] OR Tajikistan[TIAB] OR Tadzhikistan[TIAB] OR Tadjikistan[TIAB] OR Tanzania[TIAB] OR Thailand[TIAB] OR Timor-Leste[TIAB] OR East Timor[TIAB] OR Togo[TIAB] OR Togolese Republic[TIAB] OR Tonga[TIAB] OR Tunisia[TIAB] OR Turkey[TIAB] OR Turkmenistan[TIAB] OR Turkmenia[TIAB] OR Tuvalu[TIAB] OR Uganda[TIAB] OR Ukraine[TIAB] OR Uruguay[TIAB] OR Uzbekistan[TIAB] OR Vanuatu[TIAB] OR New Hebrides[TIAB] OR Venezuela[TIAB] OR Vietnam[TIAB] OR Viet Nam[TIAB] OR West Bank[TIAB] OR Gaza[TIAB] OR Yemen[TIAB] OR Zambia[TIAB] OR Zimbabwe[TIAB] OR Rhodesia[TIAB]) OR (LIC[TIAB] OR LICs[TIAB] OR MIC[TIAB] OR MICs[TIAB] OR LMIC[TIAB] OR LMICs[TIAB] OR LAMIC[TIAB] OR LAMICs[TIAB] OR LAMI countr*[TIAB])) | 24/10/2024 | 1,693,663 |
| **S4** | **S1 AND S2 AND S3** | **24/10/2024** | **1356** |
