## Supplementary material for "Management capacity of healthcare facilities in Low- and Middle-Income Countries; A scoping review": S4 File- Search terms for Web of Science

**Web of Science search strategy**

| **#** | **Search terms** | **Date searched** | **Results** |
| --- | --- | --- | --- |
| S1 | TS=("manager" OR "administrator" OR "supervisor" OR "coordinator" OR "lead" OR "overseer" OR "director" OR "executive" OR "manag*" OR "administrat*" OR "supervis*" OR "coordinat*" OR "superintendent") | 24/10/2024 | 8,600,002 |
| S2 | TS=("managerial capacity" OR "leadership capacity" OR "governance capacity" OR ("manag*" AND "capacit*") OR ("lead*" AND "capacit*") OR ("manag*" AND "competenc*") OR ("facility manag*")) | 24/10/2024 | 418,239 |
| S3 | TS=("primary health care" OR "primary care" OR "primary healthcare" OR "Primary health" OR "Community health cent*" OR "community health clinics" OR "health clinics" OR "district health" OR "preventive health service*" OR "primary level" OR "primary-level") | 24/10/2024 | 240,349 |
| S4 | TS=("primary health care" OR "primary care" OR "primary healthcare" OR "Primary health" OR "Community health cent*" OR "community health clinics" OR "health clinics" OR "district health" OR "preventive health service*" OR "primary level" OR "primary-level") AND TS=(Afghanistan OR Albania OR Algeria OR "American Samoa" OR Angola OR Antigua OR Barbuda OR Argentina OR Armenia OR Azerbaijan OR Bangladesh OR Belarus OR Belize OR Benin OR Bhutan OR Bolivia OR Bosnia OR Herzegovina OR Botswana OR Brazil OR Bulgaria OR "Burkina Faso" OR Burundi OR Cambodia OR Cameroon OR "Cape Verde" OR "Central African Republic" OR Chad OR Chile OR China OR Colombia OR Comoros OR "Costa Rica" OR "Côte d'Ivoire" OR Cuba OR Djibouti OR Dominica OR "Dominican Republic" OR Ecuador OR Egypt OR "El Salvador" OR Eritrea OR Ethiopia OR Fiji OR Gabon OR Gambia OR Georgia OR Ghana OR Grenada OR Guatemala OR Guinea OR "Guinea-Bissau" OR Guyana OR Haiti OR Honduras OR India OR Indonesia OR Iran OR Iraq OR Jamaica OR Jordan OR Kazakhstan OR Kenya OR Kiribati OR Korea OR Kosovo OR Kyrgyzstan OR Laos OR Latvia OR Lebanon OR Lesotho OR Liberia OR Libya OR Lithuania OR Madagascar OR Malawi OR Malaysia OR Maldives OR Mali OR Marshall OR Mauritania OR Mauritius OR Mexico OR Micronesia OR Moldova OR Mongolia OR Montenegro OR Morocco OR Mozambique OR Myanmar OR Namibia OR Nepal OR Nicaragua OR Niger OR Nigeria OR Pakistan OR Palau OR Palestine OR Panama OR "Papua New Guinea" OR Paraguay OR Peru OR Philippines OR Romania OR Russia OR Rwanda OR Samoa OR "São Tomé" OR Senegal OR Serbia OR Seychelles OR "Sierra Leone" OR "Solomon Islands" OR Somalia OR "South Africa" OR "Sri Lanka" OR Sudan OR Suriname OR Swaziland OR Syria OR Tajikistan OR Tanzania OR Thailand OR "Timor-Leste" OR Togo OR Tonga OR Tunisia OR Turkey OR Turkmenistan OR Tuvalu OR Uganda OR Ukraine OR Uruguay OR Uzbekistan OR Vanuatu OR Venezuela OR Vietnam OR "West Bank" OR Gaza OR Yemen OR Zambia OR Zimbabwe OR LIC OR LICs OR MIC OR MICs OR LMIC OR LMICs OR LAMIC OR lamins OR "LAMI countr*") | 24/10/2024 | 33,675 |
| **S5** | **S1 AND S2 AND S3 AND S4** | **24/10/2024** | **1283** |
