## Supplementary material for "Management capacity of healthcare facilities in Low- and Middle-Income Countries; A scoping review": S5 File- Search terms for Scopus

**Scopus search strategy**

| **#** | **Search Terms** | **Date searched** | **Results** |
| --- | --- | --- | --- |
| S1 | TITLE-ABS-KEY ( manag* AND ( capacity OR compete* ) ) | 24/10/2024 | 411,468 |
| S2 | ( ( primary OR community OR frontline OR district OR zonal OR family ) AND ( healthcare AND facility ) ) | 24/10/2024 | 174,775 |
| S3 | ( EXCLUDE ( AFFILCOUNTRY , "United States" ) OR EXCLUDE ( AFFILCOUNTRY , "United Kingdom" ) OR EXCLUDE ( AFFILCOUNTRY , "Australia" ) OR EXCLUDE ( AFFILCOUNTRY , "Canada" ) OR EXCLUDE ( AFFILCOUNTRY , "Undefined" ) OR EXCLUDE ( AFFILCOUNTRY , "Netherlands" ) OR EXCLUDE ( AFFILCOUNTRY , "Sweden" ) OR EXCLUDE ( AFFILCOUNTRY , "Switzerland" ) OR EXCLUDE ( AFFILCOUNTRY , "Belgium" ) OR EXCLUDE ( AFFILCOUNTRY , "Germany" ) OR EXCLUDE ( AFFILCOUNTRY , "Norway" ) OR EXCLUDE ( AFFILCOUNTRY , "France" ) OR EXCLUDE ( AFFILCOUNTRY , "Italy" ) OR EXCLUDE ( AFFILCOUNTRY , "Denmark" ) OR EXCLUDE ( AFFILCOUNTRY , "Spain" ) OR EXCLUDE ( AFFILCOUNTRY , "Ireland" ) OR EXCLUDE ( AFFILCOUNTRY , "Japan" ) OR EXCLUDE ( AFFILCOUNTRY , "United Arab Emirates" ) OR EXCLUDE ( AFFILCOUNTRY , "Turkey" ) OR EXCLUDE ( AFFILCOUNTRY , "New Zealand" ) OR EXCLUDE ( AFFILCOUNTRY , "Israel" ) OR EXCLUDE ( AFFILCOUNTRY , "South Korea" ) OR EXCLUDE ( AFFILCOUNTRY , "Greece" ) OR EXCLUDE ( AFFILCOUNTRY , "Austria" ) OR EXCLUDE ( AFFILCOUNTRY , "Slovakia" ) OR EXCLUDE ( AFFILCOUNTRY , "Portugal" ) OR EXCLUDE ( AFFILCOUNTRY , "Poland" ) OR EXCLUDE ( AFFILCOUNTRY , "Estonia" ) ) AND ( LIMIT-TO ( DOCTYPE , "ar" ) ) AND ( LIMIT-TO ( PUBSTAGE , "final" ) ) AND ( LIMIT-TO ( LANGUAGE , "English" ) ) |  |  |
| **S4** | **S1 AND S2 AND S3** | **24/10/2024** | **1,016** |
