## Supplementary material for "Management capacity of healthcare facilities in Low- and Middle-Income Countries; A scoping review": S6 File- Search terms for Google Scholar

**Google Scholar Search**

| **#** | **Search Terms** | **Date Searched** | **Results** |
| --- | --- | --- | --- |
| **S1** | "managerial capacity" OR "leadership capacity" OR "administrative capacity" OR "governance capacity" OR "management competence" OR "capacity building" AND ("primary health care" OR "community health" OR "health clinics" OR "district health services" OR "family medicine") | 24/10/2024 | 19,800  Picked 1^st^ 200 (Explanation in manuscript) |
